# High transmissibility of COVID-19 near symptom onset

**DOI:** 10.1101/2020.03.18.20034561

**Authors:** Hao-Yuan Cheng, Shu-Wan Jian, Ding-Ping Liu, Ta-Chou Ng, Wan-Ting Huang, for Taiwan COVID-19 outbreak investigation team, Hsien-Ho Lin

## Abstract

**Background:** The dynamics of coronavirus disease 2019 (COVID-19) transmissibility after symptom onset remains unknown.

**Methods:** We conducted a prospective case-ascertained study on laboratory-confirmed COVID-19 cases and their contacts. Secondary clinical attack rate (considering symptomatic cases only) was analyzed for different exposure windows after symptom onset of index cases and for different exposure settings.

**Results:** Thirty-two confirmed patients were enrolled and 12 paired data (index-secondary cases) were identified among the 1,043 contacts. The secondary clinical attack rate was 0.9% (95% CI 0.5–1.7%). The attack rate was higher among those whose exposure to index cases started within five days of symptom onset (2.4%, 95% CI 1.1–4.5%) than those who were exposed later (zero case from 605 close contacts, 95% CI 0–0.61%). The attack rate was also higher among household contacts (13.6%, 95% CI 4.7–29.5%) and non-household family contacts (8.5%, 95% CI 2.4–20.3%) than that in healthcare or other settings. The higher secondary clinical attack rate for contacts near symptom onset remained when the analysis was restricted to household and family contacts. There was a trend of increasing attack rate with the age of contacts (*p* for trend < 0.001).

**Conclusions:** High transmissibility of COVID-19 near symptom onset suggests that finding and isolating symptomatic patients alone may not suffice to contain the epidemic, and more generalized social distancing measures are required. Rapid reduction of transmissibility over time implies that prolonged hospitalization of mild cases might not be necessary in large epidemics.

## Main text

### Introduction

The coronavirus disease 2019 (COVID-19) outbreak originated from Wuhan has spread to more than 100 countries only two months after the virus, Severe Acute Respiratory Syndrome Coronavirus 2 (SARS-CoV-2), was identified in January 2020.^1,2^ Following the Wuhan lockdown and other extreme social distancing measures conducted by the Chinese government, several countries with massive outbreaks also implemented similar measures, including shutting down the entire cities or communities, banning international or domestic travel, conducting border control with symptom screening, and implementing isolation and quarantine to slow down the epidemic.

However, for a novel pathogen like SARS-CoV-2, its unknown epidemiologic characteristics and transmission dynamics complicated the development and evaluation of effective control policies.^3^ Researches on COVID-19 have sprouted with growing epidemics in different countries and provided some valuable insights. The short transmission cycle (serial interval) of COVID-19 and results from viral shedding studies suggested the possibility of transmission near or even before symptom onset, while prolonged viral shedding raised concerns about prolonged infectiousness and the need for extended hospital stay.^4-6^ A few preliminary contact-tracing studies showed that the high-risk exposure setting of COVID-19 transmission was in the household.^7-9^ Nevertheless, these fragmented knowledges were still inadequate to answer some practical questions like, when and how long we should isolate a COVID-19 patient or quarantine close contacts. To connect these insights and reveal the full picture of COVID-19 transmission, evidence from the field is urgently needed to provide information about the transmission risk at different time points after symptom onset and at different exposure settings.

Taiwan’s first COVID-19 case was confirmed on January 21.^10^ With proactive containment efforts and comprehensive contact tracing, the number of COVID-19 cases in Taiwan was maintained low compared to other countries overwhelmed by massive outbreaks.^11^ Using the contact tracing data in Taiwan, we aimed to delineate the transmission dynamics of COVID-19, evaluate the infection risk at different exposure windows, and estimate the infectious period.

## Methods

### Study population

In response to the outbreak in Wuhan, Taiwan Centers for Disease Control (Taiwan CDC) made COVID-19 as a notifiable disease on January 15, 2020. A prospective case-ascertained study enrolling all confirmed cases and their close contacts was conducted between January 15 and February 26.

### Ascertainment of cases

A confirmed case should meet the criteria of notification for COVID-19 in Taiwan and be tested positive by real-time reverse transcriptase-polymerase chain reaction (RT-PCR) test.^12^ Detailed information including demographic and clinical data was reported to the National Notifiable Disease Surveillance System.^13^ The investigation team determined the clinical severity of the confirmed patients following the World Health Organization (WHO) interim guidance.^14^

### Contact tracing for COVID-19

When the patient was laboratory-confirmed to have SARS-CoV-2 infection, a thorough epidemiological investigation including contact tracing was implemented by the outbreak investigation team of Taiwan CDC and local health authorities. The definition of a close contact was a person who did not wear appropriate personal protection equipment (PPE) while having face-to-face contact with a confirmed case for more than 15 minutes after symptom onset. A contact was listed as a household contact if he/she lived in the same household with the index case, while those listed as family contacts were family members not living in the same household. For health care settings, close contact was defined by contacting an index case within two meters without appropriate PPE. Medical staff, hospital workers, and other patients in the same setting were included.

All close contacts were quarantined at home for 14 days after their last exposure to the index case. During the quarantine period, any relevant symptoms (fever, cough, or other respiratory symptoms) of close contacts would trigger RT-PCR testing for COVID-19. For high-risk populations, including household and hospital contacts, RT-PCR was performed regardless of symptoms. In Taiwan CDC, we used an electronic tracing system (Infectious Disease Contact Tracing Platform and Management System) to follow and record the daily health status of those quarantined contacts. The information collected included age, gender, the index case, date of exposure, and the exposure setting.

### Data processing and analysis

Paired data of index case and close contacts were extracted from the contact tracing database and outbreak investigation reports. For a family cluster, the index case was determined based on the temporality of symptom onset and review of the epidemiological link. A secondary case was excluded from the paired data if the date of onset was earlier than the date of exposure. Similarly, a close contact would be excluded if the date of last exposure was earlier than the date of symptom onset of the index case. For health care contacts, the date of exposure would be the date of admission if the exact date of exposure was not recorded.

Incubation period and serial interval were estimated using the contact tracing data and publicly available datasets (Supplement Appendix). We used the Bayesian hierarchical model to increase the stability in small-sample estimation. The exposure window period was defined as the time period from the first contact after symptom onset of the index case to the end of contact. For household contacts who lived with the index case, the day of first contact was set to be the day of symptom onset of the index case. Secondary clinical attack rate was calculated by dividing the number of symptom confirmed cases by the number of close contacts. We analyzed the dynamic change of secondary clinical attack rate after symptom onset in several ways. First, we categorized contacts based on the time of the initial exposure to the index case after index case’s symptom onset (days 0–3, 4–5, 6–7, 8–9, or >9). Second, we considered the whole exposure window period and fitted logistic regression models to estimate the piecewise period-specific odds ratio under different definitions of exposure (binary exposure or cumulative person-time exposure in each categorical period). Third, we estimated the density function of transmission at different timing of exposure since symptom onset, considering the whole exposure window period and assuming a gamma distribution of the density function (See Supplementary appendix for details of the statistical analysis).

The data management and analysis were done using R software (R Foundation for Statistical Computing) and RStan (Stan Development Team).

## Results

By February 26, there were 32 laboratory-confirmed COVID-19 patients, including five household/family clusters and four asymptomatic patients. Sixteen (50%) patients were imported, and the remaining 16 cases were locally acquired. Of the 16 locally-acquired cases, three patients without any travel history were detected because of pneumonia of unknown etiology. The remaining 13 cases were secondary cases found through contact tracing. One of the 13 cases was excluded from subsequent transmission pair analysis because the documented day of exposure occurred after symptom onset of the secondary case. The median delay from symptom onset to confirmed diagnosis was 5 days (range 0–12) among imported cases and 18.5 days (range 2–28) among locally-acquired cases (Figure 1A). All secondary cases had their first day of exposure within five days of the index case’s symptom onset (Figure 1B). The presumed transmission trees were depicted in Figure 1C. We estimated that the mean incubation period was 4.9 days (95% credible interval [CrI] 2.7–8.4), and the mean serial interval was 7.0 days (95% CrI 3.7–13.2).

**Figure 1.**
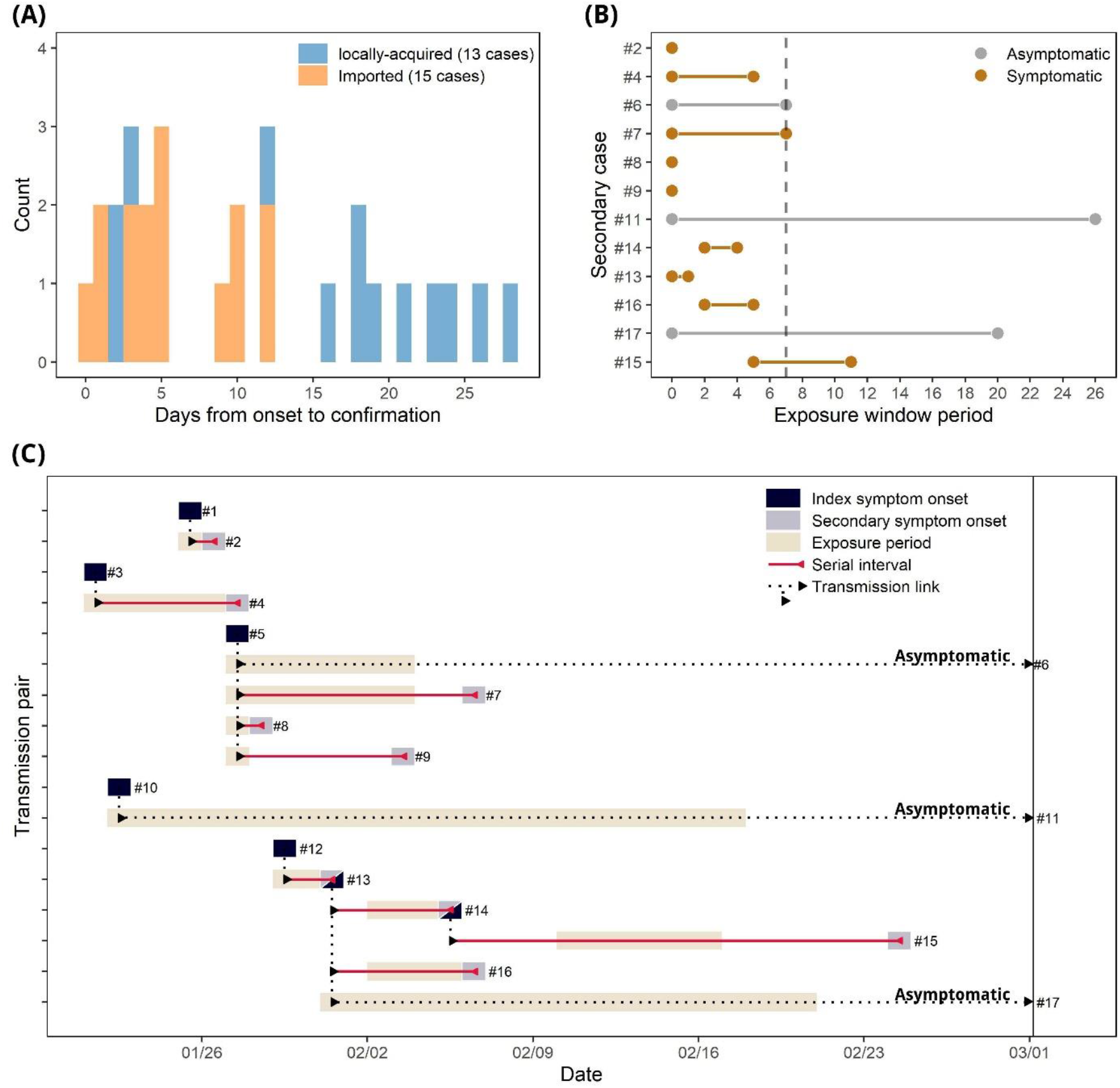
Epidemiology of confirmed COVID-19 cases in Taiwan. (A) Distribution of the interval between symptom onset to confirmed diagnosis of COVID-19 among 28 cases (excluding four asymptomatic cases). (B) Exposure window period of the 12 locally transmitted cases, defined as the time period from the first day of exposure after symptom onset of the index case to the last day of exposure. (C) Transmission tree of 12 locally transmitted pairs.

A total of 1,043 close contacts were identified. Among them, 3.4% were household contacts, 4.5% were non-household family contacts, and 28.9% were health care contacts (Table 1). The risk for COVID-19 infection (considering 12 transmitted cases) in the contacts was 1.2% (95% confidence interval [CI] 0.6–2.0%), and the secondary clinical attack rate was 0.9% (95% CI 0.4–1.6%) among all contacts. The clinical attack rate was higher among those whose initial exposure to the index case was within five days of symptom onset (2.4%, 95% CI 1.1–4.5%) than those who were exposed later (zero transmission out of 605 contacts, 95% CI 0–0.61%) (Table 2 and Figure 2A). The attack rate from early exposure remained high when we restricted the analysis to household and non-household family contacts (Table 3 and Figure 2B). Further analyses that accounted for the whole exposure window period also revealed higher transmissibility near symptom onset (Figure 2C–2D).

**Table 1.** The characteristics of close contacts by different exposure settings.

|  | Household<br>(n=36) | Family<br>(n=47) | Health care<br>(n=301) | Others<br>(n=659) |
| --- | --- | --- | --- | --- |
| **Age (median, range)** | 44 (7–96) | 49.5 (12–88) | 33 (16–87) | 39 (10–80) |
| **Age group** |  |  |  |  |
| 0–19 | 7 (19%) | 4 (9%) | 2 (1%) | 57 (9%) |
| 20–39 | 8 (22%) | 10 (21%) | 190 (63%) | 254 (39%) |
| 40–59 | 10 (28%) | 17 (36%) | 72 (24%) | 251 (38%) |
| ≥ 60 | 8 (22%) | 7 (15%) | 20 (7%) | 47 (7%) |
| Unknown | 3 (8%) | 9 (19%) | 17 (6%) | 50 (8%) |
| **Sex** |  |  |  |  |
| Male | 17 (47%) | 22 (47%) | 88 (29%) | 366 (56%) |
| Female | 19 (53%) | 25 (53%) | 204 (68%) | 260 (39%) |
| Unknown | 0 (0%) | 0 (0%) | 9 (3%) | 33 (5%) |
| **Time from onset to exposure\***<br>**(median, range)** | 0 (0–5) | 1. (0–26) | 8 (0–23) | 7 (0–26) |
| **Time from onset to exposure\*** |  |  |  |  |
| ≤ 3 days | 35 (97%) | 10 (21%) | 45 (15%) | 209 (32%) |
| 4–5 days | 1 (3%) | 0 (0%) | 14 (5%) | 59 (9%) |
| 6–7 days | 0 (0%) | 9 (19%) | 14 (5%) | 61 (9%) |
| 8–9 days | 0 (0%) | 3 (6%) | 104 (35%) | 276 (42%) |
| > 9 days | 0 (0%) | 22 (47%) | 123 (41%) | 10 (2%) |
| Unknown | 0 (0%) | 3 (6%) | 1 (0%) | 44 (7%) |

\* Defined as the elapsed time between the date of symptom onset of the index case and the first date of exposure.

**Table 2.** Secondary clinical attack rate for COVID-19 among close contacts by different exposure attributes.

|  | No. of secondary cases<br>(asymptomatic case) | No. of<br>contact | Secondary clinical<br>attack rate<br>(95% CI) | Risk ratio<br>(95% CI) |
| --- | --- | --- | --- | --- |
| **Exposure setting** |  |  |  |  |
| Household* | 7 (2) | 36 | 13.9% (4.7–29.5%) | 1.00 |
| Family | 5 (1) | 47 | 6.5% (1.4–17.9%) | 0.59 (0.17–2.03) |
| Health care | 0 | 301 | 0% (0–1.2%) | 0 |
| Others | 0 | 659 | 0% (0–0.6%) | 0 |
| **Time from onset to exposure\*\*** |  |  |  |  |
| ≤ 3 days | 11 (3) | 299 | 2.7% (1.2–5.2%). | 1 |
| 4–5 days | 1 (0) | 74 | 1.4% (0–7.3%) | 0.50 (0.06–3.94) |
| 6–7 days | 0 (0) | 84 | 0% (0–4.3%) | 0 |
| 8–9 days | 0 (0) | 383 | 0% (0–0.1%) | 0 |
| > 9 days | 0 (0) | 155 | 0% (0–2.4%) | 0 |
| **Age group of close contacts** |  |  |  |  |
| 0–19 | 1 (1) | 70 | 0% (0–5.1%) |  |
| 20–39 | 3 (1) | 462 | 0.4% (0.1–1.6%) | 1 |
| 40–59 | 5 (1) | 348 | 1.1% (0.3–2.9%) | 2.66 (0.49–14.41) |
| ≥ 60 | 3 (0) | 82 | 3.6% (0.8–10.3%) | 8.45 (1.43–49.8) |
| **Imported index case** |  |  |  |  |
| No | 10 (3) | 485 | 2.0% (1.0–3.8%) | 1 |
| Yes | 2 (0) | 558 | 0.4% (0–1.3%) | 0.17(0.04–0.79) |
| **Clinical severity of index case** |  |  |  |  |
| Mild illness | 4 (0) | 230 | 1.7% (0.5–4.4%) | 1.00 |
| Mild pneumonia | 2 (1) | 26 | 0.4% (0.1–2.1%) | 0.22 (0.02–1.96) |
| Severe pneumonia | 0 (0) | 277 | 0% (0–1.3%) | 0 |
| ARDS/Sepsis | 6 (2) | 275 | 1.4% (0.4–3.7%) | 0.84 (0.21–3.33) |

\*One foreign caregiver in the hospital was included as household contact.
\*\* Defined as the elapsed time between the date of symptom onset of the index case and the first date of exposure.
ARDS, acute respiratory distress syndrome.

**Table 3.**
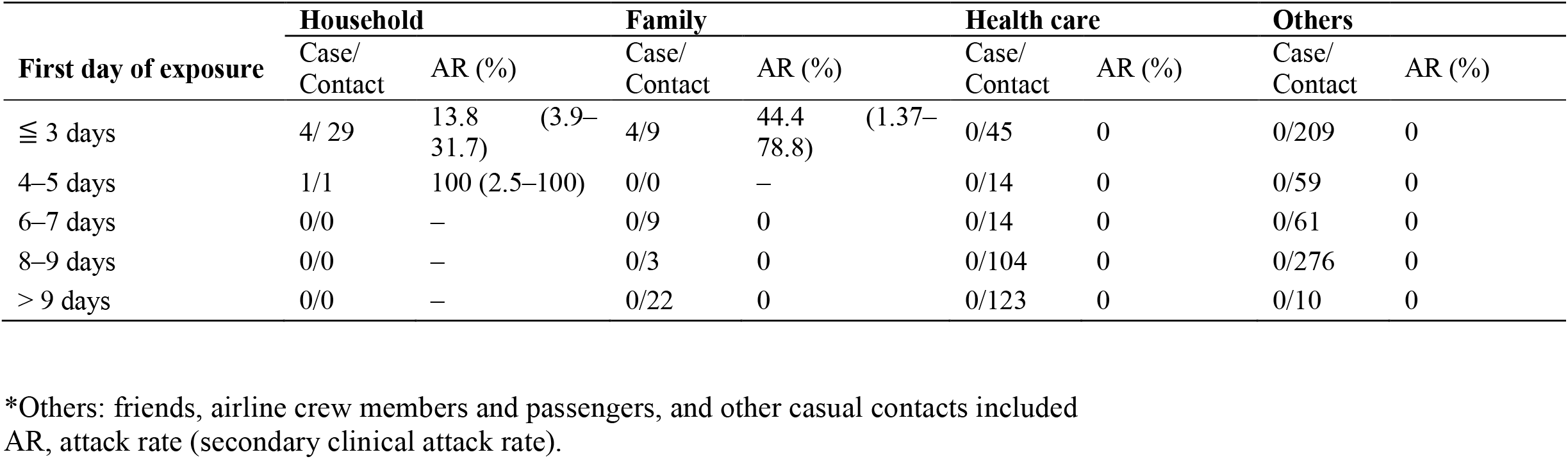
The risk for symptomatic COVID-19 infection among close contacts, simultaneously stratified by exposure setting and time from symptom onset of the index case to first day of exposure.

**Figure 2.**
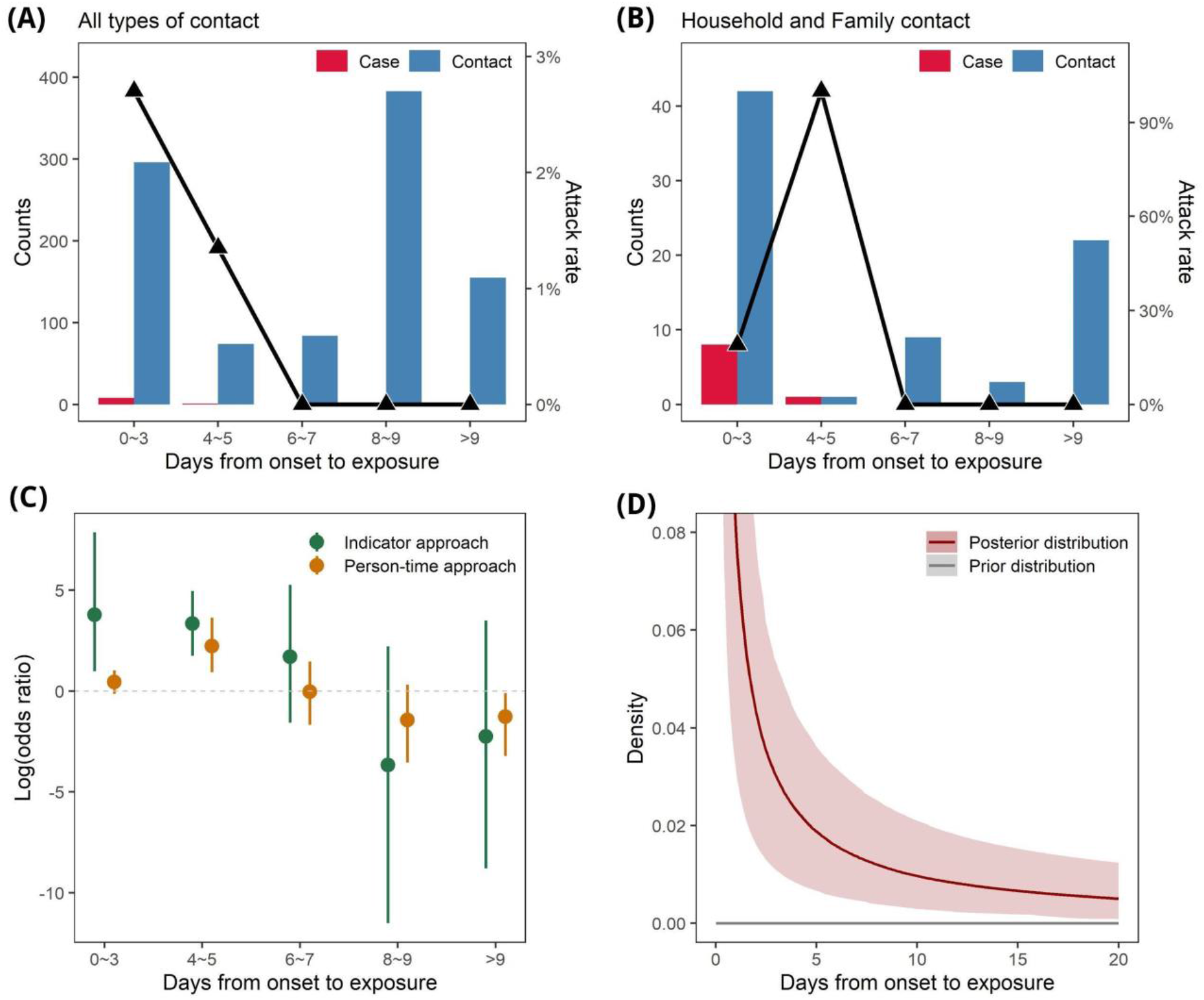
Number of close contacts and secondary symptomatic cases, and secondary clinical attack rate by the time from symptom onset of index cases to the date of first exposure (A) among all contacts and (B) household and family contacts. (C) Odds ratio (log scale) of secondary disease by the different exposure period after symptom onset; the exposure dose was defined by the indicator approach (green) and the person-time approach (orange). (D) Estimated density function of transmission over the exposure period after symptom onset. See Supplementary Appendix for details.

The secondary clinical attack rate was 13.6% (95% CI 4.7–29.5%) among household contacts and 8.5% (95% CI 2.4–20.3%) in non-household family contacts (Table 2). No nosocomial infection was observed in this study. There was a trend of increasing secondary clinical attack rate with the age of contacts, ranging from 0% (95% CI 0– 5.1%) in those aged less than 20 years to 3.6% (95% CI 0.8–10.3%) in those aged 60 years and above (*p* for linear trend: <0.001). Close contacts of confirmed cases presented with mild illness were not at a lower risk compared to the contacts of more severe cases. The secondary attack rate among contacts of locally-acquired cases was higher than that among contacts of imported cases (Table 2).

## Discussion

This study is one of the very few initial reports of the secondary clinical attack rate among close contacts to confirmed COVID-19 cases. Our analysis revealed the early transmission dynamics with relatively short infectious period of COVID-19: a higher transmission risk around the time of symptom onset of the index case, followed by a lower transmission risk at the later stage of disease. The observed decreasing transmission risk over time for COVID-19 was in striking contrast to the transmission pattern of SARS, in which the transmission risk remained low until after day 5 of symptom onset in the index cases.^15^

Our finding might explain the substantially shorter transmission cycle (serial interval) of COVID-19 than that of SARS. Following Nishiura et al,^4^ we updated the information on transmission pairs from published reports and included 48 pairs. The updated serial interval had a mean of 5.4 days (95% CrI 4.1–7.2 days) (Supplementary Appendix). In contrast, the mean serial interval of SARS was estimated to be 8.4 days in Singapore.^15^ Our analysis suggested that the shorter serial interval of COVID-19 was due to the combination of early-stage transmission and a short period of infectiousness.

The observed pattern of secondary clinical attack rate over time was also consistent with the quantitative data of the SARS-CoV-2 viral shedding in upper respiratory specimens, which reported a high viral load around the time of symptom onset, followed by a gradual decrease in viral shedding to a low level after 10 days.^5^ The viral load was similar among asymptomatic, minimally symptomatic, and symptomatic patients. Another virological study in COVID-19 patients also found no viable isolates of the virus after the first week of symptoms. Our data agreed with the virological data on high transmissibility in the first week and much decreased risk afterwards.^16^

To summarize the evidence, the decreasing risk for secondary infection over time in our study, the observed short serial interval, and the trend of decreasing viral shedding and viability after symptom onset strongly suggested high transmissibility of the disease near or even before the day of symptom onset. Since the onset of overt clinical symptoms such as fever, dyspnea, and signs of pneumonia usually occurred 5–7 days after initial symptom onset, the infection might well have been transmitted onward at the time of detection.^17,18^ This characteristic makes the containment efforts challenging. In a modeling study, Hellewell et al. found that the possibility of controlling COVID-19 through isolation and contact tracing decreased with increasing proportion of transmission that occurred before symptom onset.^19^ This simulation combined with our findings might explain the difficult situation in Wuhan, South Korea, Iran, and Italy. Aggressive social distancing and proactive contact tracing might be necessary to block the transmission chain of COVID-19 and to keep presumptive patients away from susceptible populations with a high risk for severe disease.

The observed short duration of infectiousness with lower risk of transmission one week after symptom onset had critical implications on redirecting the control efforts of COVID-19. Given the non-specific and mostly mild symptoms of COVID-19 at presentation, patients are often identified and hospitalized at the later stage of disease when the transmissibility has started to decrease. In this case, hospitalization would not be helpful for isolation and reducing transmission, and should be spared for severe patients. When the number of confirmed cases increases rapidly, home care for patients with mild illness should be considered.^20^ In Taiwan, the most prolonged duration of hospital isolation for the 32 confirmed cases was more than one month. If every patient with mild illness is to be isolated in the hospital or other isolation institutes for such a prolonged period, the healthcare system will soon be overwhelmed during a large epidemic. In this case, an unusually high case fatality rate such as that observed in Wuhan may occur.^21,22^ Similarly, better understanding of the potential duration of transmission could help direct containment strategies. For example, the efforts of contacting tracing could focus on the contacts near or even before symptom onset of the index cases when the number of index cases or contacts is too large to afford. To fight a potential pandemic like COVID-19, improved efficiency for resource allocation will be critical since a massive outbreak will rapidly consume public health and medical resources.

Several patients in our study had pneumonia with unknown etiology and had multiple contacts in the healthcare setting before being diagnosed, but none of these healthcare contacts resulted in nosocomial transmission. Besides the basic PPE used by medical staffs, this result might be due to their late admissions and lower transmissibility by the time of hospitalization as well. This pattern is compatible with the observation in China and Hong Kong. In China, the number of nosocomial infections might be lower than that reported because some healthcare workers acquired infections in their households rather than in the healthcare setting;^9^ In Hong Kong, most hospitalization was also delayed to at least 5 days after disease onset.^23^ In closed settings like hospital or cruise ship,^24,25^ fomite transmission might play an important role, amplifying the risk of transmission and making the temporality of transmission less identifiable.^25-27^ Better understandings of the dynamic change of transmissibility over time and of the transmission route of infected health care workers could further reduce the unnecessary infection control measures and over-equipped PPEs, and thus reduce healthcare workers’ workloads during large outbreaks.

Our study has limitations. First, we did not completely examine contacts before the symptom onset of the index cases. Therefore, we might have underestimated the importance of early transmission. In other words, the actual importance of early transmission could be higher than our estimates. Our findings agree with the recommendation from WHO on using four days before symptom onset as the starting date for contact tracing.^28^ This modification might further reveal the pattern of early transmission in COVID-19. Second, we could not completely separate out the effect of close household contact and early contact given the strong correlation of the two. The increased transmissibility in the early stage of COVID-19 may be partially attributed to the effect of household and non-household family contacts rather than increased infectiousness at the early stage. But the pattern of early transmission remained when we stratified by type of exposure.

In summary, our analysis suggested that the majority of transmission of COVID-19 occurred at the very early stage of the disease, and the secondary clinical attack rate among contacts decreased over the time line of symptom development. The pattern of high transmissibility near symptom onset and possible short infectious period may drastically change the thinking of control strategies for COVID-19. More studies are urged to elucidate the transmission dynamics of this novel disease clearly.

## Data Availability

The data is available upon request.

## Ethics approval

Information collection were done by the pronouncement of the Central Epidemic Command Center (CECC), and, in accordance with Article 17 of the Communicable Disease Control Act. As part of the public health response functions of the CECC for surveillance purposes, institutional review board approval of this study was waived. The data was deidentified prior to analysis.

## Acknowledgment

We thank the Taiwan COVID-19 outbreak investigation team (as listed in Supplement Appendix), and staffs of regional control centers of Taiwan CDC and public health bureaus (Taipei city, New Taipei city, Taoyuan city, Changhua county, and Kaohsiung county) for their dedicated outbreak investigation and meticulous date collection. Our works could not be done with their efforts. We also thank Chia-Lin Lee for the development of electronic contact tracing system (Epidemic Intelligence Center, Taiwan CDC); Ching-Hung Wang (Tony Q) for the consultation of system development; Angela Song-En Huang for writing assistance.

